# Predicting social distancing index during COVID-19 outbreak through online search engines trends

**DOI:** 10.1101/2020.05.28.20115816

**Authors:** P. C. Lins-Filho, M. M. S. Araújo, T. S. Macêdo, A. K. A. Ferreira, M. C. F. Melo, E. L. M. S. Silva, J. L. M. Freitas, A. F. Caldas

## Abstract

Online-available information has been considered an accessory tool to estimate epidemiology and collect data on diseases and population behavior patterns. This study aimed to explore the potential use of Google and YouTube relative search volume to predict social distancing index in Brazil during COVID-19 outbreak and verify the correlation between social distancing measures with the course of the epidemic. Data concerning the social distancing index, epidemiological data on COVID-19 in Brazil and the search engines trends for “Coronavirus” were retrieved from online databases. Multiple linear regression was performed and resulted in a statistically significant model evidencing that Google and YouTube relative search volumes are predictors of the social distancing index. The Spearman correlation test revealed a weak correlation between social distancing measures and the course of the COVID-19 epidemic. Health authorities can apply these data to define the proper timing and location for practicing appropriate risk communication strategies.

## Introduction

The World Health Organization have recently declared South America as the new coronavirus epicenter, mainly because of the situation in Brazil that registers the most cases and deaths in Latin America [1]. Given the COVID-19 pandemic, robust risk communication is urgently needed particularly in the most affected countries [2]. Internet query platforms, which allows to interact with internet-based data, have been considered a source of potentially useful and accessible resources, especially aimed to identify outbreaks and implement intervention strategies [3, 4]. Online-available information has been considered as a surrogate tool for estimating epidemiology and gathering data about patterns of disease and population behavior [5–7].

As the online queries on COVID-19 increases globally reflecting the interest of people to be aware about this emerging infectious disease, mining online data and search patterns on electronic resources might provide a better support to manage this worldwide health crisis [8]. Internet searches and social media data have been reported to correlate with traditional surveillance data and can even predict the outbreak of disease epidemics several days or weeks earlier [9]. Evidence points that Google Trends could potentially define the proper timing and location for practicing appropriate risk communication strategies for affected populations and be employed to predict outbreak trends of the novel coronavirus [2, 5, 10].

Previous investigations reported the use of Internet search engines as source of data for public health surveillance and diseases incidence prediction worldwide, as zika, in Brazil and Colombia [11]; influenza, in the United States [12]; malaria, dengue fever and chikungunya, in India [13]; and Middle East respiratory syndrome, in Korea [14]. However, there are no reports on assessing the relative search volume (RSV) of search engines to predict social distancing behavior during infectious diseases outbreaks.

Predictions might support in health resource management and planning for prevention purposes [5]. As COVID-19 treatment protocol is still uncertain it is especially important to prevent the virus dissemination in society [15]. Currently, the virus spread prevention approaches focus on hand hygiene, social distancing and quarantine [16]. Social distancing is designed to reduce interactions between people in a broader community, in which individuals may be infectious but have not yet been identified hence not yet isolated. This measure is particularly useful in settings where community transmission is believed to have occurred, but where the linkages between cases is unclear, and where restrictions placed only on persons known to have been exposed is considered insufficient to prevent further transmission [15, 17, 18]. Collective infection control measures can reduce the disease incidence, though at the price of a prolongation of the epidemic period [19]. Therefore, it is important to raise information on these measures.

A recent study that assessed the impact of online information on the individual-level intention to voluntarily self-isolate during the pandemic concluded that in order to enhance individuals’ motivation to adopt preventive measures such as social distancing, actions should focus on raising consciousness on the severity of the situation, in addition, information overload had a significant impact on individuals’ threat and coping perceptions, and through them on self-isolation intention [20]. Thus, the aim of the present investigation was to predict social distancing index (SDI) through Google and YouTube search trends and investigate the correlation between the SDI with epidemiological data on COVID-19 outbreak in Brazil.

## Methods

In Brazil, the current market share of Google among the search engines is over 97% [21]. Google Trends (https://trends.google.com/trends/) data is a randomly collected sample of Google search queries, each piece of data is categorized and tagged with a topic. Each data point is divided by the total searches in a specific location over a time period to compare relative popularity. Google Trends portrays search frequency output as a normalized data series and the resulting numbers are scaled on a range of 0 to 100 based on a topic proportion to all searches on all topics. Scores represents search interest relative to the highest point on the graph for that time period and geographic location. A value of 100 is the peak popularity of a term. A value of 50 indicates that the term is half as popular as it was at its peak of popularity [22]. This same methodology was applied to YouTube search trends. To the present investigation the Brazilian Portuguese correspondent topic for “coronavirus”, which held most popularity, was used.

The Social Distancing Index was created to help combat the spread of COVID-19, since its launch it has been improved with the sole objective of providing an increasingly accurate data for public authorities and research institutes. In order to achieve the index, highly accurate geolocation data was treated with a distance algorithm. Polygons from all regions of the Brazilian Institute of Geography and Statistics were adopted in order to ensure a more accurate categorization and more reliable data [23]. Data is available on Inloco website, displayed as map and chart.

Epidemiological data concerning COVID-19 outbreak in Brazil were collected from the Brazilian government Health Ministry database, available online [24]. Statistical data on daily new cases, cumulative number of cases, cumulative number of deaths and recovered cases were retrieved.

All databases were assessed for data collection on 23 May 2020, and the information corresponds to the period from 23 February to 20 May 2020.

Data were submitted to statistical analysis, all tests were applied considering an error of 5% and the confidence interval of 95%, and the analyzes were carried out using SPSS software version 23.0 (SPSS Inc. Chicago, IL, USA). Although the hypothesis of normal distribution of data was not confirmed by the Kolmogorov-Smirnov test, the statistical analysis was performed by the application of nonparametric tests. The strength of the association between distinct measures was tested with Spearman rank correlation. Multiple linear regression was performed to verify whether Google and YouTube relative search volumes are predictors of the social distancing index in Brazil.

## Results

The multiple linear regression analysis resulted in a statistically significant model [F (2,85) = 32,045; p<0.001; R2 = 0.430]. Therefore, Google RSV (β = 1.226; t = 7.887; p<0.001) and YouTube RSV (β = –0.930; t = –5.980; p<0.001) are predictors of the social distancing index in Brazil. The equation that describes this relationship is (SDI) = 34.347 + 0.422 (Google RSV) + (−0.359) (YouTube RSV).

In Brazil for the time span analyzed the mean SDI score was approximately 43%, the maximum of social distancing observed during this period was 62.2%. In mean scores over 3312 new cases of COVID-19 were confirmed daily. In the moment of data collection, the reported total number of cumulative deaths, confirmed cases and recovered cases were 18859, 291579 and 116683, respectively. The mean values of search engines RSV are shown in table 1.

**Table 1.** Frequency measures of SDI, search engines RSV and COVID-19 daily new cases for the evaluated period.

|  | Mean <sup>(SD)</sup> | Median | Minimum | Maximum |
| --- | --- | --- | --- | --- |
| Social distancing index | 43.126 <sup>(8.130)</sup> | 43.30 | 26.60 | 62.20 |
| Google RSV | 37.470 <sup>(23.649)</sup> | 34.50 | 2 | 100 |
| YouTube RSV | 19.520 <sup>(21.035)</sup> | 11.50 | 2 | 100 |
| Daily new cases | 3312.26 <sup>(4564.24)</sup> | 1180 | 0 | 19951 |

\*RSV – Relative Search Volume

Correlation between SDI and the other studied variables was found to be varying from weak to moderate, statistically significant correlation was found with all measures tested except for cumulative recovered cases, as shown in table 2.

**Table 2.**
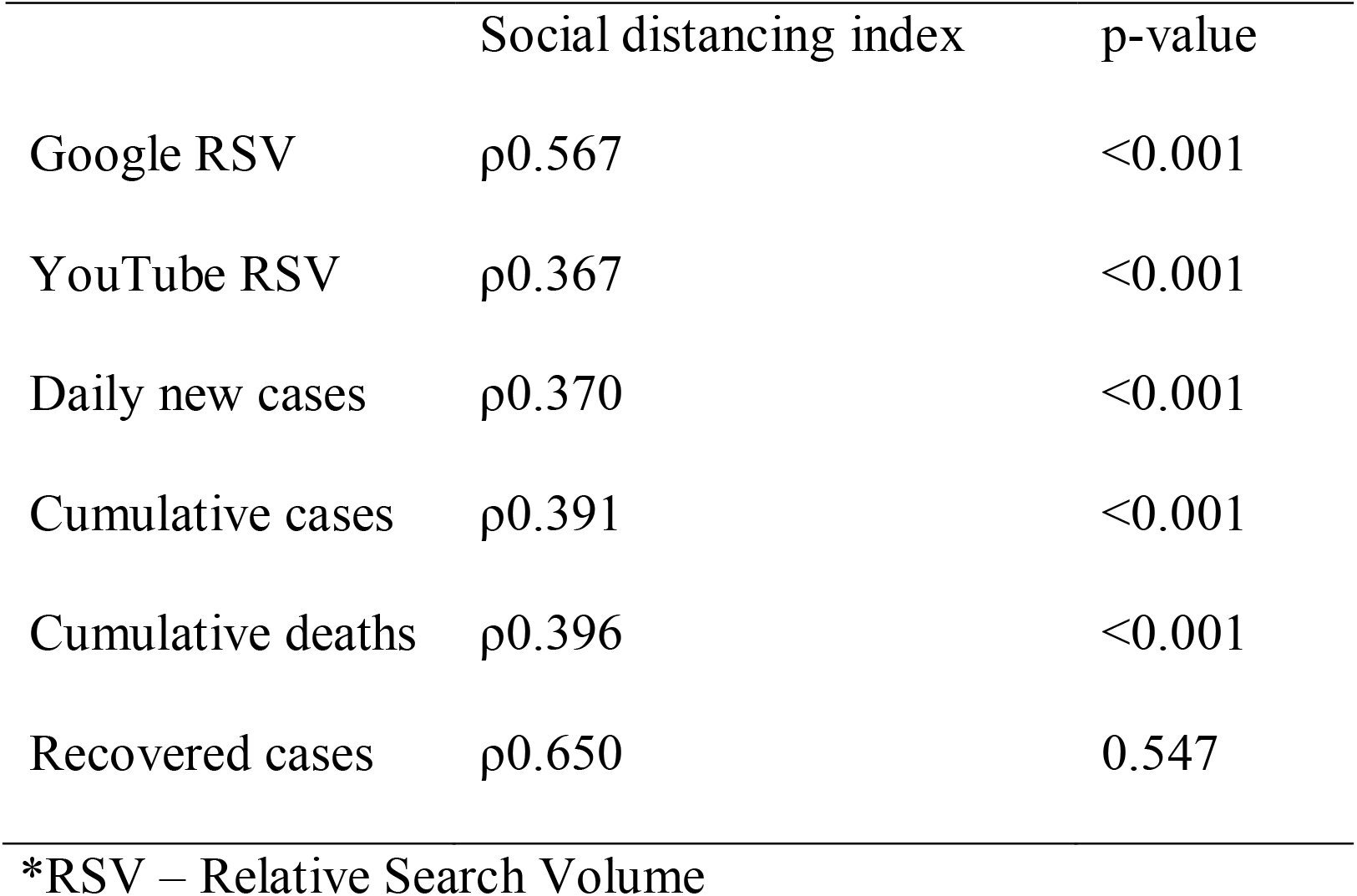
Correlation between SDI measures, COVID-19 epidemiological data and search engines RSV.

## Discussion

Evidence suggests that collective isolation measures have been highly effective in controlling the spread of the COVID-19 [16, 25]. However, maintaining isolation for many months may have even worse consequences than an epidemic wave that runs an acute course, the isolation measures should be thoughtfully planned and executed based on current stage of pandemic [26]. As observed in table 1 the mean score for SDI was 43.126 with a discrete standard deviation when compared with the standard deviation of the daily new cases. A weak correlation was found between the isolation measure with epidemiological data from COVID-19, this may represent that in Brazil social isolation measures are poorly associated with the course of the disease in the country. In addition, these findings may be correlated with failure to control the increase in cases that were added daily, on average, by 3312.26 new cases, during the period covered by this investigation. The absence of concise social distancing policies and, furthermore, the political instability at the center of the Brazilian government poses a deadly distraction in the middle of a public health emergency [1].

The weak correlation between social distancing and the course of the disease can also be observed in the time series pattern seen in figure 1 (section A), where while the number of cases per day and cumulative deaths show an ascending pattern, the social distancing index remains with slight changes, with the exception of a slight increase at the end of March, when cases start to show an ascending profile. At the end of the time series, when the daily new cases are at peak, SDI was under de mean observed in the evaluated period.

**Figure 1.**
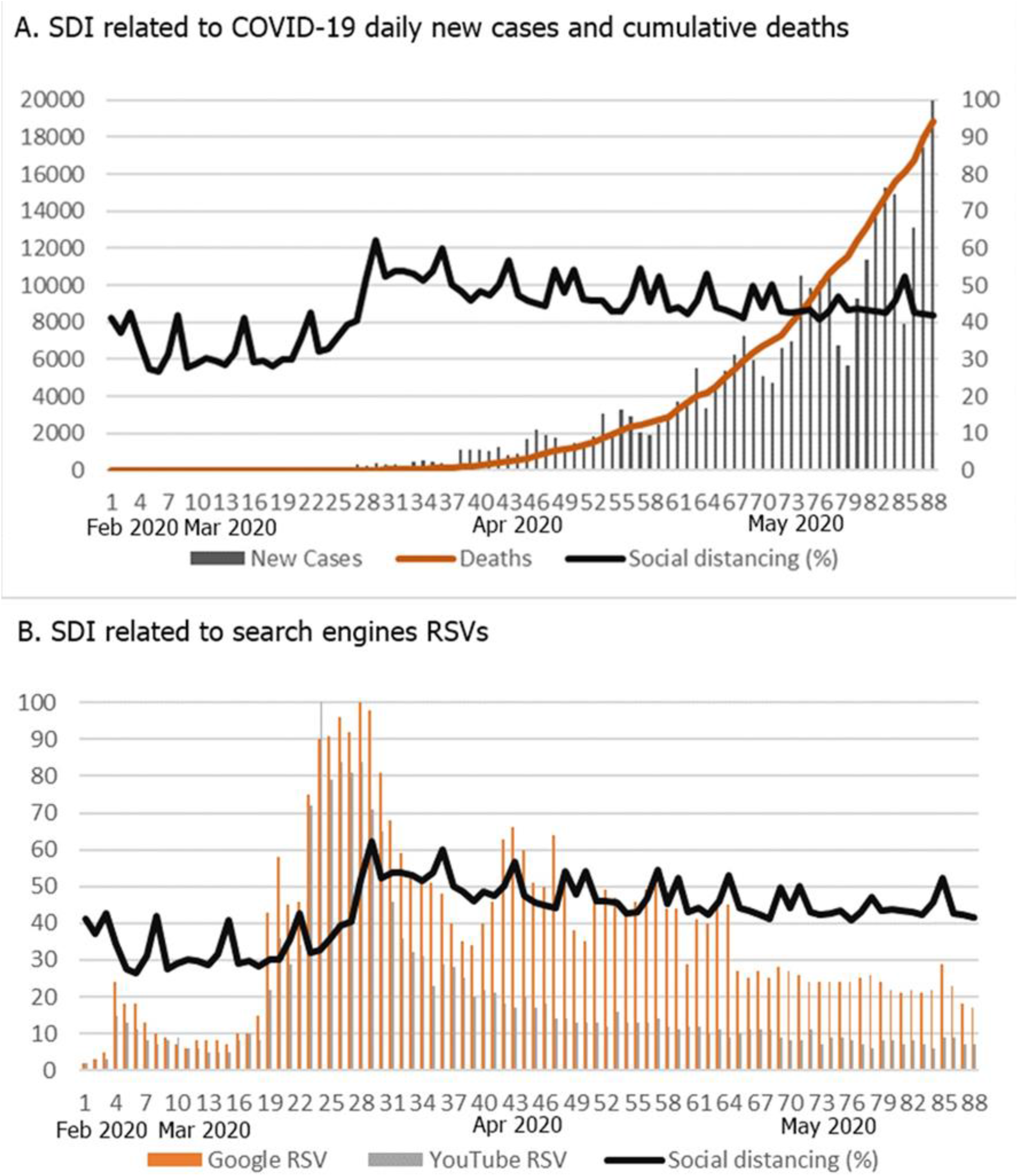
Time series of social distancing index related to Google and YouTube RSVs and COVID-19 new cases and cumulative deaths in Brazil.

A correlation was found between search engines RSV and SDI (table 2). To further investigate this correlation a multiple linear regression was performed, data extracted from this analysis showed that Google RSV and YouTube RSV are predictors to social distancing. In figure 1 (section B) it is possible to observe de matching behavior of this measures along the time series, with a similar pattern of peaks and decrease starting at the end of April.

The positive correlation found with Google RSV may be associated with the access to information, since raising awareness on the severity of the situation and the importance on following the advice from health organizations is a key point on achieving self-isolation intention [20].

The negative association observed between YouTube RSV and SDI through de multiple linear regression may correlate with the low quality of YouTube content on COVID-19 reported in previous studies [27–29], since misinformation can hinder the communication of health professionals and organizations with general public and even reduce compliance with treatments or medical advices [30, 31].

The findings of the present study support the evidence that online-available information can potentially assist conventional epidemiologic tools for estimating data about patterns of disease and population behavior [5, 6]. Relative search volume of Google and YouTube could define the proper timing and location for practicing appropriate risk communication strategies. Health authorities might apply these data to measure the effect of the transmission of information on the population and to obtain feedback from research statistics.

## Data Availability

The data used in this study are available.

## Acknowledgments

P.C., T.M., M.A. and E.L. were supported by a PhD scholarship from Coordenação de Aperfeiçoamento de Pessoal de Nível Superior (CAPES).

## References

(1) Anon. COVID-19 in Brazil: “So what?”[Editorial]. The Lancet 2020; 395: 1461.

(2) Husnayain A, Fuad A, Chia-Yu ES. Applications of Google Search Trends for Risk Communication in Infectious Disease Management: A Case Study of the COVID-19 Outbreak in Taiwan. International journal of infectious diseases: IJID: official publication of the International Society for Infectious Diseases 2020; 95: 221–223.

(3) Barros JM, Duggan J, Rebholz-Schuhmann D. The Application of Internet-Based Sources for Public Health Surveillance (Infoveillance): Systematic Review, Journal of Medical Internet Research 2020; 22: e13680.

(4) Bhattacharya S. Predicting emerging and re-emerging disease outbreaks through internet search trends: An analysis from India. AIMS Public Health 2019; 6: 1–3.

(5) Ayyoubzadeh SM, et al. Predicting COVID-19 Incidence Through Analysis of Google Trends Data in Iran: Data Mining and Deep Learning Pilot Study. JMIR public health and surveillance 2020; 6: e18828.

(6) Walker A, Hopkins C, Surda P. The Use of Google Trends to Investigate the Loss of Smell Related Searches During COVID-19 Outbreak. International forum of allergy & rhinology 2020.

(7) Cervellin G, Comelli I, Lippi G. Is Google Trends a reliable tool for digital epidemiology? Insights from different clinical settings. Journal of Epidemiology and Global Health 2017; 7: 185‐189.

(8) Effenberger M, et al. Association of the COVID-19 pandemic with Internet Search Volumes: A Google Trends^TM^ Analysis. International Journal of Infectious Diseases 2020; 95:192‐197.

(9) Ortiz-Martínez Y, et al. Can Google® trends predict COVID-19 incidence and help preparedness? The situation in Colombia. Travel Medicine and Infectious Disease 2020.

(10) Higgins TS, et al. Correlations of Online Search Engine Trends With Coronavirus Disease (COVID-19) Incidence: Infodemiology Study. JMIR Public Health and surveillance 2020; 6: e19702.

(11) Morsy S, et al. Prediction of Zika-confirmed Cases in Brazil and Colombia Using Google Trends. Epidemiology and infection 2018; 146: 1625–1627.

(12) Pei S, et al. Forecasting the Spatial Transmission of Influenza in the United States. Proceedings of the National Academy of Sciences of the United States of America 2018; 115: 2752–2757.

(13) Verma M, et al. Google Search Trends Predicting Disease Outbreaks: An Analysis from India. Healthcare Informatics Research 2018; 24: 300–308.

(14) Shin S-Y, et al. High correlation of Middle East respiratory syndrome spread with Google search and Twitter trends in Korea. Scientific Reports 2016; 6: 1–7.

(15) Guner R, HasanogLu I, Aktas F. COVID-19: Prevention and control measures in community. Turkish Journal of Medical Sciences 2020; 50: 571–577.

(16) Taghrir MH, Akbarialiabad H, Marzaleh MA. Efficacy of Mass Quarantine as Leverage of Health System Governance During COVID-19 Outbreak: A Mini Policy Review. Archives of Iranian medicine 2020; 23: 265–267.

(17) Wilder-Smith A, Chiew CJ, Lee VJ. Can we contain the COVID-19 outbreak with the same measures as for SARS?. The Lancet Infectious Diseases 2020; 20: e102–e107.

(18) Wilder-Smith A, Freedman DO. Isolation, quarantine, social distancing and community containment: pivotal role for old-style public health measures in the novel coronavirus (2019-nCoV) outbreak. Journal of Travel Medicine 2020; 27: taaa020.

(19) Raoult D, et al. Coronavirus infections: Epidemiological, clinical and immunological features and hypotheses. Cell Stress 2020; 4: 66–74.

(20) Faroog A, Laato S, Islam AKMN. Impact of Online Information on Self-Isolation Intention During the COVID-19 Pandemic: Cross-Sectional Study. Journal of medical Internet research 2020; 22: e19128.

(21) Statcounter Search Engine Market Share Brazil See (https://gs.statcounter.com/search-engine-market-share/all/brazil). Accessed 20 April 2020.

(22) Mavragani A, et al. Assessing the Methods, Tools, and Statistical Approaches in Google Trends Research: Systematic Review. J Med Internet Res 2018; 20: e270.

(23) Inloco Mapa brasileiro da COVID-19 (https://mapabrasileirodacovid.inloco.com.br/pt/). Accessed 23 May 2020.

(24) Brazil Painel Coronavírus (https://covid.saude.gov.br/). Accessed 23 May 2020.

(25) Sjördin H, et al. Only Strict Quarantine Measures Can Curb the Coronavirus Disease (COVID-19) Outbreak in Italy, 2020. Eurosurveillance 2020; 25: 2000280.

(26) Ioannidis, JPA. Coronavirus disease 2019: The harms of exaggerated information and non‐evidence‐based measures. European Journal of Clinical Investigation 2020, e13223.

(27) Basch CH, et al. Preventive Behaviors Conveyed on YouTube to Mitigate Transmission of COVID-19: Cross-Sectional Study. JMIR public health and surveillance 2020; 6: e18807.

(28) Basch CE, et al. The Role of YouTube and the Entertainment Industry in Saving Lives by Educating and Mobilizing the Public to Adopt Behaviors for Community Mitigation of COVID-19: Successive Sampling Design Study. JMIR public health and surveillance 2020; 6: e19145.

(29) Li HOY, et al. YouTube as a source of information on COVID-19: a pandemic of misinformation? BMJ Global Health 2020; 5: e002604.

(30) Lu X, et al. Relationship Between Internet Health Information and Patient Compliance Based on Trust: Empirical Study. Journal of medical Internet research 2018; 20: e253.

(31) Lu X, Zhang R. Impact of Physician-Patient Communication in Online Health Communities on Patient Compliance: Cross-Sectional Questionnaire Study. Journal of medical Internet research 2019; 21: e12891.

